# The Importance of Measuring Context in the Study of Whole Person Health: Healing and Persistence of Clinical Effect in Acupuncture Science

**DOI:** 10.1101/2025.05.13.25327395

**Authors:** Lisa Conboy, Xiaoming Sheng

## Abstract

The last 50 years we have seen an increase in published biomedical science on acupuncture, how it works and for which conditions. Our analysis considers if persistence of treatment effect may be related to continued treatment after study completion. Our data comes from a completed Army-funded Randomized Controlled Trial “The Effectiveness of Acupuncture in the Treatment of Gulf War Illness (Army grant W81XWH-09-2-0064)”. This pragmatic RCT applied acupuncture treatment in a therapeutically informed dose determined in focus groups with expert practitioners. We followed the advice of the practitioners and designed a treatment protocol of Individualized acupuncture treatments administered by practitioners in the community for a possible treatment window of 6 months. Using data from our focus groups we designed the study to compare the dose that our acupuncture focus group experts told us would be necessary for effect (twice per week) to once per week treatment. Treatment protocols individualized or personalized to the patient are standard of care in acupuncture therapy. Put another way, a standardized treatment protocol was not used; instead experienced licensed practitioners were given training in the known medical information of GWI, and encouraged to treat with discretion while keeping detailed treatment records. The Importance of Measuring Context to Study Whole Person Health: Healing and Persistence of Clinical Effect in Acupuncture Science

## Introduction

The last 50 years we have seen an increase in published biomedical science on acupuncture, how it works and for which conditions^i^. Acupuncture has been incorporated into many health delivery systems^ii^ and the results of such are arguably positive, such as Medicare coverage for acupuncture treatments for chronic Low Back Pain. Yet there remains a lack of consensus on how to model acupuncture as a treatment. Should acupuncture be conceptualized as a drug, with dose-response effects? Or is acupuncture treatment more of a behavioral intervention as it requires action on the part of the patient?

Acupuncture is a complex intervention that appears to be highly influenced by contextual elements such as the quality of the patient-practitioner relationship, and the patient’s expectations for treatment^iii,iv.^ While many acupuncturists think of behavior modification as part of their tool kit^v^, and some biomedical research models acupuncture as a behavioral intervention^vi^, more commonly acupuncture is modeled like a drug treatment; more acupuncture is a larger dose than less acupuncture, and larger doses will have greater effects on most patients than smaller doses. Further if a patient stops taking their “drug” be it a pharmaceutical pain reliever, or acupuncture treatments, the symptoms of illness will often return.

Randomized Controlled Trials (RCTs) are considered the gold standard of medical research^vii^. RCTs are powerful, particularly for investigations in which the mechanism of action is known and controllable. Knowledge of the mechanism of action is necessary to create an appropriate control group to eliminate (as much as possible) the influence of context so that we can securely look at the effects of a single mechanisms of action. RCTs implement this control with techniques such as randomization to treatment groups, and using tight inclusion criteria, with the goal of eliminating competing explanations of any seen changes in the study outcomes. As biomedicine focuses on biological changes in the body, the health influence of the patients’ context is often considered less important^viii^. The strength of the RCT design to control for contextual factors further facilitates this idea that contextual factors can be partitioned while we study true effects on the biological level. More recently assumption has changed in some subfields of medical science. For example quality metrics such as the Revised STandards for Reporting Interventions in Clinical Trials of Acupuncture (STRICTA)^ix^, now encourage the recording of contextual factors in clinical trials, which will help remedy our current avoidance of the influence of context.

There is high quality evidence from meta-analyses that in the treatment of pain, the effect of acupuncture care persists long after active study treatment has ended. MacPherson and colleagues reviewed 20 RCTs which included 6376 patients, testing the clinical effect of acupuncture in the treatment of chronic pain conditions including musculoskeletal pain (low back, neck, and shoulder), osteoarthritis of the knee, and headache/migraine. Using mean estimate scores, approximately 90% of the clinical benefit of acupuncture relative to controls was maintained at 12 month follow up^x^. However the trials that went into this review did not offer data on subjects’ health behavior after trial completion. Perhaps some subjects continued to use acupuncture and this high persistence of effect is actually due to continued treatment.

Our analysis considers if persistence of treatment effect may be related to continued treatment after study completion. Our data comes from a completed Army-funded Randomized Controlled Trial “The Effectiveness of Acupuncture in the Treatment of Gulf War Illness (Army grant W81XWH-09-2-0064)”. This pragmatic RCT applied acupuncture treatment in a therapeutically informed dose determined in focus groups with expert practitioners^xi^. We followed the advice of the practitioners and designed a treatment protocol of *Individualized* acupuncture treatments administered by practitioners in the community for a possible treatment window of 6 months.

Using data from our focus groups we designed the study to compare the dose that our acupuncture focus group experts told us would be necessary for effect (twice per week) to once per week treatment. Treatment protocols individualized or personalized to the patient are standard of care in acupuncture therapy^xii^. Put another way, a standardized treatment protocol was not used; instead experienced licensed practitioners were given training in the known medical information of GWI, and encouraged to treat with discretion while keeping detailed treatment records.

In this original analysis we found a statistically significant effect of acupuncture on the symptoms of pain and physical function, and significant differences in the effect of once versus twice per week treatment^xiii^. In order to explore persistence of treatment effect, subjects were resurveyed 5 years later using the same survey (Army grant DOD W81XWH-15-1-0695) as the parent trial. Here we report on the results of that resurvey, and focus on the same outcome as the MacPherson pain persistence analysis^ix^.

## Methods

All subjects from the parent trial (n=104) were invited to participate in the resurvey. To support a healthy response rate, subjects were invited to participate in the resurvey up to three times. The SurveyMonkey© platform was used to collect responses.

The SPSS© data analysis system was used for description and analysis. Missing data were not replaced. We first considered how representative the resurveyed sample was of the parent sample. Using the Independent-Samples Kolmogorov-Smirnov Test we looked to see if the parent and resurveyed sample had significantly different mean pain levels at various measurement time points in the parent trial. We next considered if use of acupuncture after the trial ended was related to level of pain relief. We used general linear models to compare difference in McGill pain scores between different treatment groups and number of treatments after the trial ended.

## Results

Fifty-one subjects from the original sample responded to the resurvey. Table 1 considers if the group that was resurveyed had a different distribution of McGill pain scores compared to the original sample. This is a way to consider if our resurvey sample was different from the full parent sample.

**Table 1:** Displayed are comparisons of mean McGill pain scores in the group that was resurveyed, those that were not resurveyed, and the full sample. The final column lists the calculated probability, and tells us that the resurveyed and not resurveyed samples are statistically different to p<0.05.

| Measurement time point | Resurveyed Subsample Mean (SD) N | Not Resurveyed Subsample Mean (SD) N | Full Sample Mean (SD) N | Independent-Samples Kolmogorov-Smirnov Test significance |
| --- | --- | --- | --- | --- |
| Baseline | 28.5 (7.6) 48 | 31.0 ( 9.4) 47 | 29.7 (8.6) 95 | 0.22 |
| 2 months | 29.4 (7.1) 48 | 31.5 (9.2) 39 | 30.3 (8.1) 87 | 0.61 |
| 4 months | 27.8 (8.3) 46 | 28.3 (8.7) 37 | 28.0 (8.4) 83 | 0.97 |
| 6 months | 26.6 (8.2) 47 | 28.9(9.1) 35 | 27.6 (8.6) 82 | 0.75 |
| Long-term follow up | 28.2 (9.3) 51 | N/A | 28.2 (9.3) 51 |  |

Using the Independent-Samples Kolmogorov-Smirnov Test, there was no significant difference between the distribution of McGill pain scores in the group that was resurveyed at follow up vs the group that were not resurveyed. Thus, at least on the variable of pain, those that were resurveyed were not different from the original full parent trial sample.

Next we considered the *persistence of treatment effect*. Looking again at Table 1, we can calculate the percent of improvement that persisted by looking at results for the full sample in the fourth column. Mean baseline pain score was 29.7 which fell only moderately to 27.6 at 6 months and 28.2 at long-term follow-up. This is an overall change baseline to 6 months of 2.1 points (29.7-27.6). Baseline to long term follow-up shows an overall change of 1.5 points (29.7-28.2=1.5), which suggests that 71% of the clinical effect was maintained (1.5/2.1).

As interesting, if we consider the only the part of the sample that was resurveyed (column 2), only 16% of the pain improvement persists (28.5-26.6=1.9 point change baseline to 6 months; 28.5-28.2= 0.3 point change baseline to long-term follow up; 0.3/1.9=0.16). This suggests that the resurvey sample may be different in terms of long term effect from the full parent trial sample.

It is rational to consider if subjects had used acupuncture after the trial, and if so whether they had used acupuncture recently. Table 2 shows the breakdown of use since the trial (in the last 5 years). One participant did not answer this question, therefore excluded from our analyses below.

**Table 2:** Frequency of use of acupuncture on resurvey.

|  | Frequency | Percent |
| --- | --- | --- |
| Less than 1 year ago | 8 | 16 |
| 1 to 3 years ago | 20 | 40 |
| More than or equal to 4 years ago | 22 | 44 |
| Total | 50 | 100 |

Recent acupuncture treatment could offer symptom relief that looks like persistence of treatment effect from the original trial, so next we removed that part (n=8) of the sample that reported using acupuncture in the last year. Of those that had **not** used acupuncture in more than 3 years (n=22), we compared persistence of effect by what treatment group. We can see that more of the treatment effect was maintained in the group that received the therapeutic dose that our acupuncture therapist told us would be necessary (2tx/ week group, see Figure A) suggesting that the degree of persistence of treatment effect in the sample was related to dose of acupuncture received. The theoretically supported dose of 2 treatments per week, that we were given by our practitioners in focus group, had significantly greater maintenance of treatment gains even years after treatment had ended.

**Figure A:**
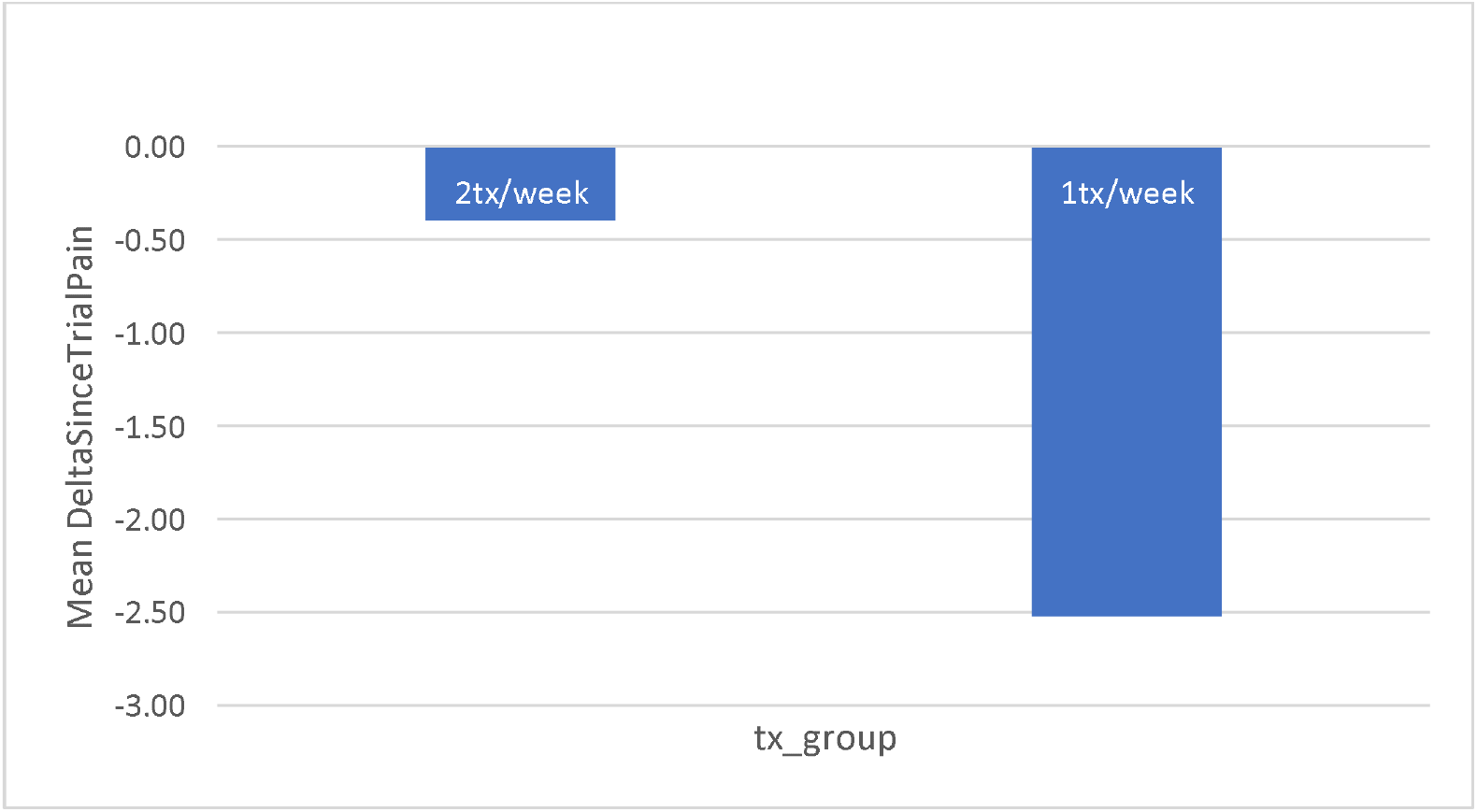
Comparing maintenance of gains by differences doses of acupuncture. **All subjects in this subsample had not received acupuncture in the last 3 years**.

These two groups are not significantly different however, p=0.255 when using an independent samples t-test (t=1.18). That is, while we can see a trend in Figure A, the results are not statistically significant to p<0.05. This could be because of small sample size, or that there is a great deal of variability in the sample of those that had not received acupuncture in the last 3 years (figure B).

**Figure B:**
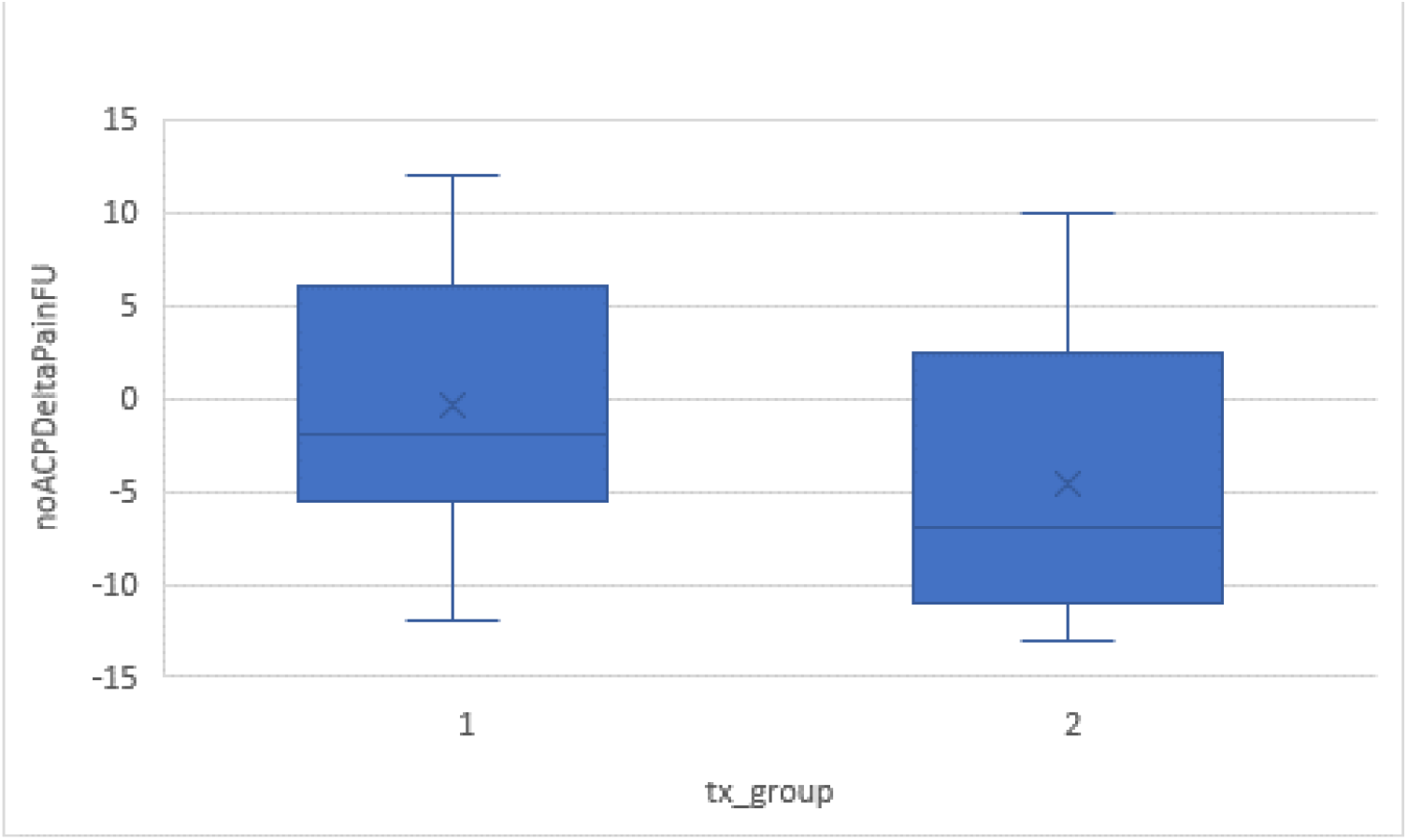
Box plot of change per dose level for those individuals who did not continue acupuncture treatment post trial. Note the large variability around the central mean. (group 1 = “2tx/week”, group 2 = “1tx/week”)

Next we looked at subjects that had acupuncture treatments after the trial ended and within the last 3 years (n=20). We first considered if the number of acupuncture treatments since the trial ended was related to maintenance of gains seen in the original trial. Figure C suggests that the number of treatments post-trial is related to treatment group during the trial, with the twice per week group (higher dose) having a higher number of treatments post study. A non-parametric Mann-Whitney rank sum test showed a marginally difference in number of treatments with an asymptotic P=0.12.

**Figure C:**
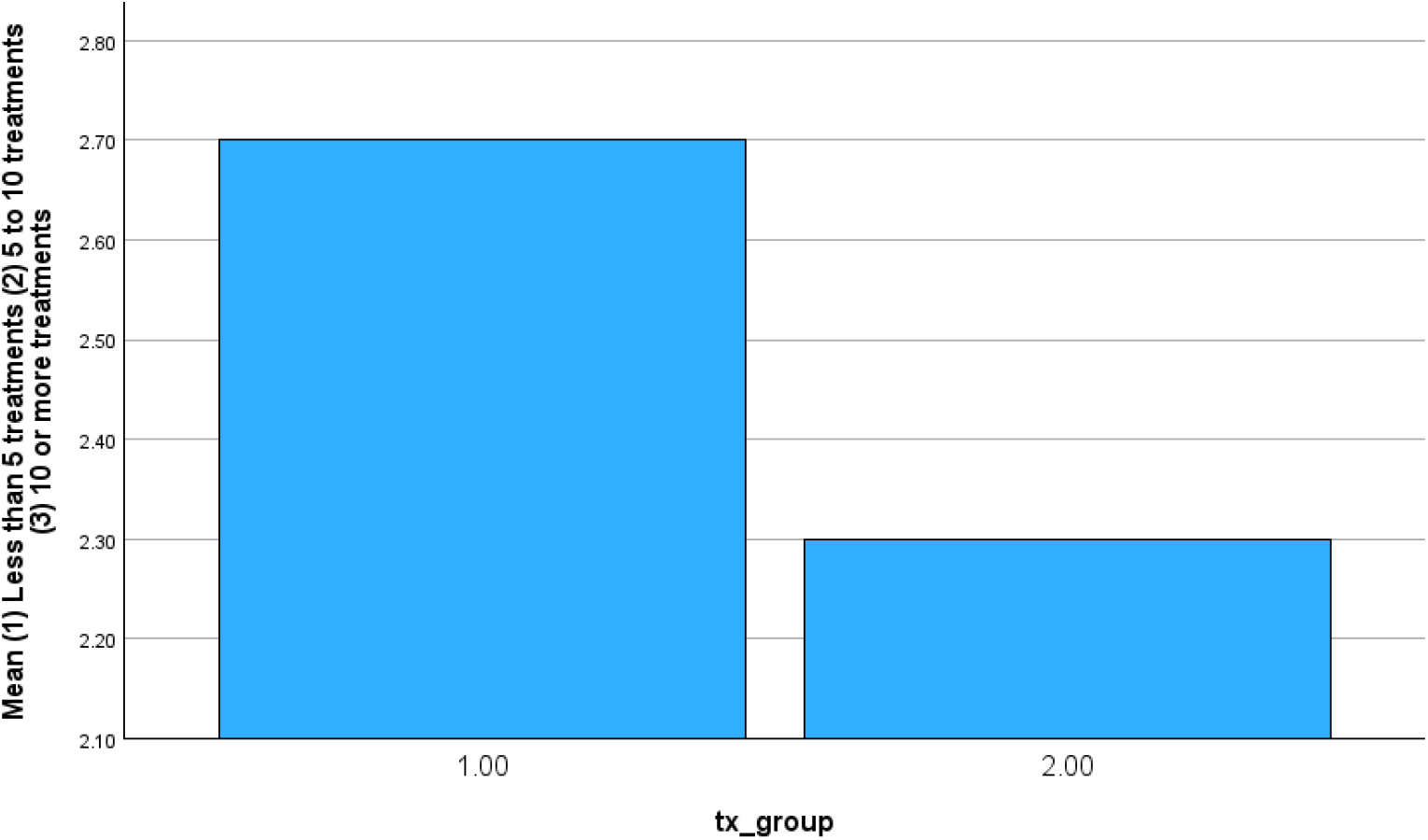
Number of treatments post-trial by treatment group. (group 1 = “2tx/week”, group 2 = “1tx/week”)

As the higher dose treatment group on average had a better effect during the trial, the choice to use more acupuncture after the trial is likely related to amount of relief found during the trial.

Next we looked at direct relationship of pain to number of treatments post-trial, adjusting for treatment group in the parent trial. This way we can probably tell whether pain is mainly due to treatment during the trial, or post-trial treatment (as the two are somewhat “correlated” with each other), or contributed by both, either independently or combined. We fit a general linear model to accommodate the ordinal nature of number of treatments.

Figure D is a plot of mean McGill pain measure by number of treatments post-trial and treatment group at the parent trial. It showed the lines crossed each other, implying the trend are different in the two treatment groups. General linear model showed their interaction has a significant P=0.031 (Table 3). In treatment group 1 (“2 tx per week”), as number of treatments post-trial increases, pain level decreases and then the effect stays; however, in treatment group 2 (“1 tx per week”), participants pain adversely increases. A limitation here is the small sample size, any significant difference could be due to chance.

**Table 3.** General Linear Model of McGill Pain at Follow Up.

| Source | Sum of Squares | d.f. | Mean Square | F | P-value |
| --- | --- | --- | --- | --- | --- |
| Number of treatments | 78.678 | 1 | 78.678 | 1.248 | .280 |
| Tx Group | 301.019 | 1 | 301.019 | 4.776 | .044 |
| Interaction | 352.980 | 1 | 352.980 | 5.601 | .031 |
| Error | 1008.341 | 16 | 63.021 |  |  |
| Total | 1440.200 | 19 |  |  |  |

**Figure D.**
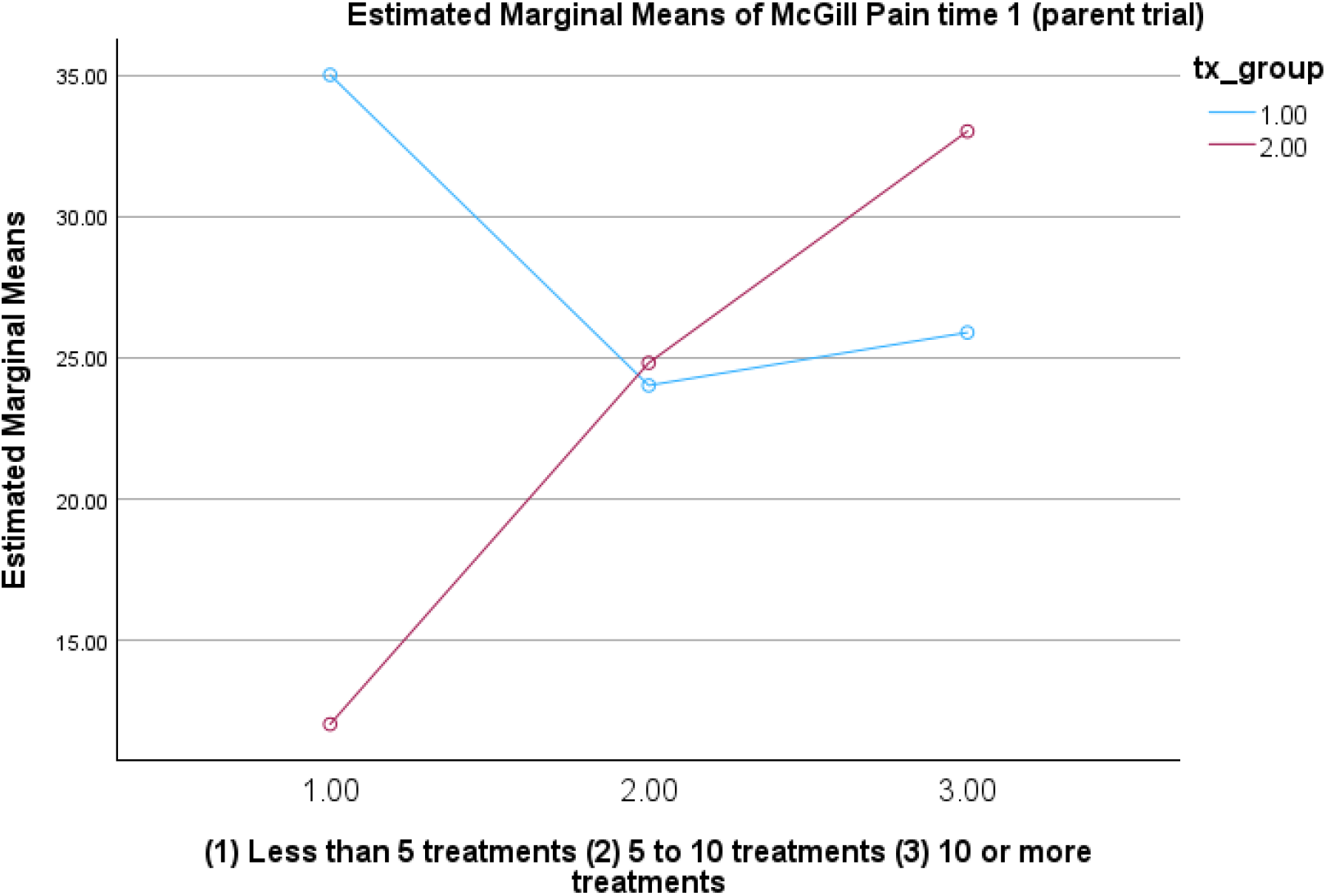
Mean of McGill Pain by number of treatments and intervention group (group 1 = “2tx/week”, treatment group 2 = “1tx/week”)

## Discussion

Data from high quality systematic reviews indicate that acupuncture treatment effects persist in the case of pain. But this effect is determined using studies that did not consider post-study use of acupuncture. Research including longer term outcomes will allow us to see persistence and variance in real world context. This is important information for persistence has ramifications for patient care and ultimate cost effectiveness.

In our small sample of an exclusively veteran population we did find some persistence of effect, but this could be explained by continued use of acupuncture after the study period. Clearly however, those in the study group that received the therapeutic dose of two treatments per week were more likely to use more acupuncture after the trial had ended.

As interesting is that treatment group is related to later use of acupuncture is the interaction found between treatment group and use of acupuncture after the study. Perhaps the group that received the therapeutic dose (the 2x per week group) had enough of a dose to get to a new stable state that requires less maintenance. The 1x per week subjects never had enough treatment to get to a new state of health. The stable state idea fits with the trend seen in the 1x per week group, with more post treatment acupuncture being related to more pain. In this group pain level may be more of a marker of how bad their (not yet sufficiently treated) pain is, and thus we see more severe cases seeking more acupuncture post-trial.

### Limitations and next steps

This analysis was done on a small sample of a population that may not be representative of the US population. Veterans may access continued acupuncture care more frequently than others because acupuncture is available to them in many VA hospitals. Still, this analysis suggests that although the RCT, and reviews of RCTs, are the strongest methods we have, if contextual elements of treatment are not considered then results may be difficult to interpret.

Further we don’t know the type of acupuncture treatment received post study. We did not ask if veterans stayed with their same practitioner or switched to a different provider. Also while effective, VA treatments may not be individualized as the treatments were in our original study. While this may seem trivial, our study indicates that type of acupuncture, and dose of treatment, are important in amount of relief achieved and maintained.

Future work could also include a more precise measure of treatments received after the study; here we used a categorical variable. It would also be useful to learn what other treatments and self-care were used after the trial. We found that those with good results during the trial were more likely to continue with acupuncture. Perhaps good results with other treatments will encourage use of other types of remedies and self-care.

## Data Availability

All data produced in the present study are available upon reasonable request to the authors

